# Minimizing Population Health Loss in Times of Scarce Surgical Capacity

**DOI:** 10.1101/2020.07.26.20157040

**Authors:** Benjamin Gravesteijn, Eline Krijkamp, Jan Busschbach, Geert Geleijnse, Isabel Retel Helmrich, Sophie Bruinsma, Céline van Lint, Ernest van Veen, Ewout Steyerberg, Kees Verhoef, Jan van Saase, Hester Lingsma, Rob Baatenburg de Jong, collaborators

**Author notes:** Both authors contributed equally (Benjamin Gravesteijn, Eline Krijkamp). Value Based Operation Room Triage team collaborators: Chris Bangma, Ivo Beetz, Patrick Bindels, Alexandra Brandt-Kerkhof, Danielle van Diepen, Clemens Dirven, Tjebbe Galema, Jeanette Goudzwaard, Mieke Hazes, Sjoerd Lagarde, Harmke Polinder-Bos, Eva Maria Roes, Hanneke Takkenberg, Mark van Vledder.

## Abstract

**Background:** COVID-19 has put unprecedented pressure on healthcare systems worldwide, leading to a reduction of the available healthcare capacity. Our objective was to develop a decision model that supports prioritization of care from a utilitarian perspective, which is to minimize population health loss.

**Methods:** A cohort state-transition model was developed and applied to 43 semi-elective non-paediatric surgeries commonly performed in academic hospitals. Scenarios of delaying surgery from two weeks were compared with delaying up to one year, and no surgery at all. Model parameters were based on registries, scientific literature, and the World Health Organization global burden of disease study. For each surgery, the urgency was estimated as the average expected loss of Quality-Adjusted Life-Years (QALYs) per month.

**Results:** Given the best available evidence, the two most urgent surgeries were bypass surgery for Fontaine III/IV peripheral arterial disease (0.23 QALY loss/month, 95%-CI: 0.09-0.24) and transaortic valve implantation (0.15 QALY loss/month, 95%-CI: 0.09-0.24). The two least urgent surgeries were placing a shunt for dialysis (0.01, 95%-CI: 0.005-0.01) and thyroid carcinoma resection (0.01, 95%-CI: 0.01-0.02): these surgeries were associated with a limited amount of health lost on the waiting list.

**Conclusion:** Expected health loss due to surgical delay can be objectively calculated with our decision model based on best available evidence, which can guide prioritization of surgeries to minimize population health loss in times of scarcity. This tool should yet be placed in the context of different ethical perspectives and combined with capacity management tools to facilitate large-scale implementation.

**Summary box:** *What is already known on this topic:* The perspective of maximizing population health, a utilitarian ethical perspective, has been described to be most defendable in times of scarcity. To prioritize surgical patients, literature mainly discusses approaches which are intra-disciplinary (e.g. within gynecological or oncological surgery) and mostly existed of narrative reviews of the literature. Some decision tools were developed, which rely on the consensus of experts on various measures of urgency (e.g. health benefit, or time until inoperable). No approach was found which transparently weighs objective factors in order to quantify a clinically relevant measure of urgency.

*What this study adds:* In contrast to previously developed approaches, our approach transparently and consistently aggregates best available objective evidence across disciplines. This novel aggregated urgency measure can be easily linked with capacity management tools. Our approach can help to minimize health losses when trying to overcome delay in surgeries in times of surgical scarcity, during the COVID-19 pandemic and beyond.

## Background

COVID-19 has put unprecedented pressure on healthcare systems worldwide. The healthcare demand of this pandemic supersedes available healthcare capacity, far beyond the demand that was imposed by the 2017 influenza pandemic.^1,2^ The pressure on the available healthcare capacity impacts the continuity of regular care. Among other things because 1) wards and operating theaters are converted to COVID-19 care facilities,^3^ 2) physicians are deployed to care for COVID-19 patients,^4,5^ and 3) the fear of contagion with SARS-CoV-2 may leave non-COVID patients reluctant to seek care^4,5^, as was seen in similar health crises like the SARS epidemic.^6^

Delay in surgical care may dramatically impact health care quality and accessibility. In the first weeks of the COVID-19 crisis in the Netherlands, 75-90% fewer surgeries were performed compared to previous years.^7^ The delay in cancer surgery already has made a large impact in the life expectancy of oncological patients.^8^ Moreover, it may be impossible to treat the whole accumulating group of patients, as estimated for orthopaedic and cardiothoracic surgery in the US.^9,10^ Because of these problems, hospitals are facing a dilemma: Which patients should be prioritized?

As stated by Emanuel et al., “*The question is not whether to set priorities, but how to do so ethically and consistently, rather than basing decisions on individual institutions’ approaches or a clinician’s intuition in the heat of the moment*”.^2^ In practice, individual surgical patients are most often triaged by experts from the respective surgical fields.^11^ Unfortunately the level of agreement on prioritization between experts is low.^12^ Additionally, prioritization across disciplines is complicated by the high degree of specialization in modern medicine. Most importantly, this approach does not systematically optimize population health. While the perspective of maximizing population health, a utilitarian ethical perspective,^13^ has been described to be most defendable in times of scarcity. ^2,14–18^

To guide prioritization of semi-elective surgeries across disciplines from a utilitarian perspective, our study aims to develop a decision model to estimate the impact of postponing surgery on health.

## Methods

### Overview

The most frequently performed semi-elective^i^ surgeries in our institute were selected. Data about these surgeries were collected and used in a broadly applicable computer-based model to estimate the effect of surgical delay on survival and health related quality of life (QoL).

### Patients and setting

The evaluated surgeries in this study comprised of non-paediatric and non-obstetric, semi-elective^i^ surgeries in Erasmus University Medical Center, an academic tertiary referring hospital in the Netherlands. From the electronic patient registry (ChipSoft, HiX), the number of surgeries, surgery time, length of stay at an intensive care unit (ICU), and length of stay at a non-ICU of all non-urgent surgeries were retrieved from July 2017 to December 2019. Next, two senior clinicians selected the semi-elective surgeries from this list. Finally, the Value Based operation room (OR) team collaborators approved the selection. Ultimately, 43 semi-elective surgeries were selected that were performed more than 80 times during the inclusion interval. Where relevant, mild and severe cases undergoing the surgery were distinguished based on clinical insight of our collaborators.

#### Input parameters

The model required 7 input parameters: 1) survival rates pre-surgery, 2) survival rate post-surgery, 3) QoL pre-surgery, 4) QoL post-surgery, 5) mean age of patients undergoing the surgery, 6) time until no effect of treatment can be expected on survival or 7) time until no effect of treatment can be expected on QoL (see Appendix A). The class of collected evidence was defined as class I (Randomized Controlled Trials (RCT) or systematic reviews of RCTs), class IIa (Prospective observational studies, before-after studies), class IIb (Retrospective observational studies, expert panels for the utilities, national registries), and class III (expert opinion).

### Markov model

A three-state cohort state-transition model was developed. This model simulates a hypothetical cohort of patients over a defined period in fixed time intervals, called cycles, to estimate the average time individuals spend in the various health conditions, called health states.^19,20^ Individuals could transition between a preoperative state, a postoperative state, and a dead state (Figure 1). Based on the time spent in these states, health benefits, like life years (LYs) or quality-adjusted life-years (QALYs) are calculated.^20–22^ The entire cohort started in the preoperative state, and was followed their remaining lifespan, until they were 100 years old, using weekly cycles. The transition from the preoperative state to the postoperative state was set to a specific week, depending on the scenario. Scenarios of surgical delay of two weeks were compared with surgical delay up to surgical delay of a year using intervals of ten weeks. In addition, the scenario where patients never received treatment was evaluated: this was modelled by following patients their entire remaining lifespan in the preoperative health state. In all scenarios, the transitions from the pre- and postoperative states to the dead state were based on survival data. A description of the model parameters and assumptions can be found in Appendix C.

**Figure 1.**
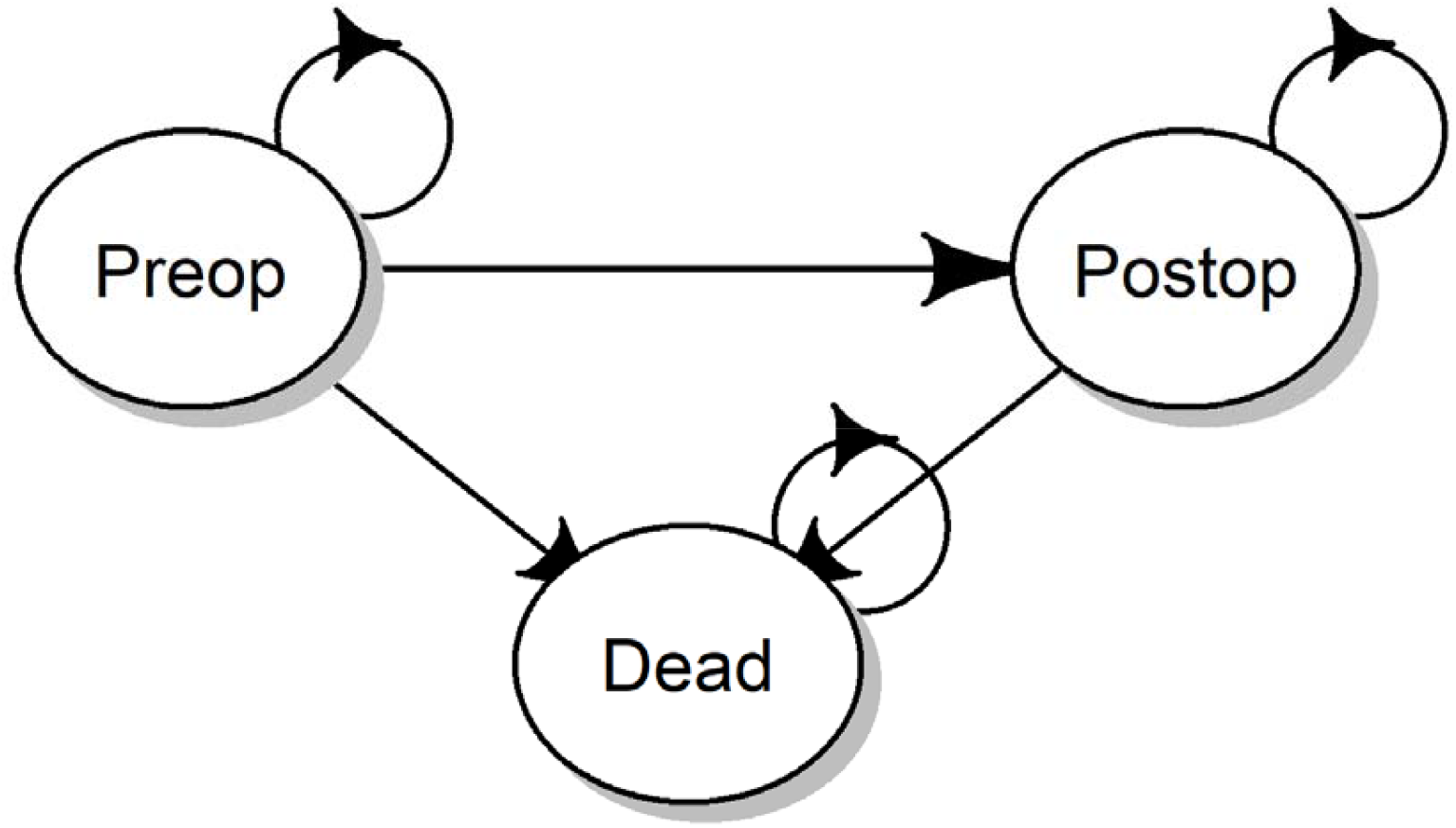
state-transition diagram of the cohort model. The model is a state transition model with three health states, a preoperatieve health states (Preop), a postoperatieve state (Postop) and Dead. All patients start in the Preop health states. This is the health states where patient eligible for surgery start in our simulation. We follow these patients over time using fixed time intervals of 1 week, these fixed time intervals are called cycles. Every cycle, patients can transition to one of the other health states or they can remain in the health states they currently are. From the Preop state they either die (transition to dead state) or continue to wait for their surgery (stay in the Preop state, the arrow points back into the health state). At the time of surgery, which is determined by us, all individuals still alive in the Preop health state transition to the Postop health state. The remaining lifetime the cohort is followed. They can die (transition from the Postop state to Dead state) or stay alive in the Postop health state (transition back to the Postop state). Finally, patients in the Dead state remain dead, so every cycle they stay in the dead state.

#### Health Effects of Surgery

The effects of delays in surgery on the health outcomes (LYs and QALYs) were evaluated. LYs disregard QoL (QoL = 100%), while QALYs incorporate QoL and are therefore preferred. The health outcomes without surgery were compared to the health outcomes with surgery at 2 weeks and 52 weeks to determine the overall health outcomes associated with surgery and health outcomes lost per 50 weeks. This measure of urgency was converted to loss per month (loss/month) and was used to rank the surgeries, where a high loss/month indicates an urgent surgery.

#### Analysis

Probabilistic sensitivity analysis (PSA) was used to incorporate parameter uncertainty in the model outcome (Appendix C). Rankings based on health benefits or health loss per unit of time were compared using Spearman’s rank correlation coefficient.

The available surgical resources should also be optimally used. Therefore, the model results were compared visually to the capacity requirements in our hospital, obtained from the electronic patient registry. Since OR room availability was the bottleneck in our hospital during the first COVID-19 wave, this study focused on surgery time.

The model was built with R software^23^ and adapted from previously published code.^24,25^ The full model code is available on GitHub: https://github.com/bgravesteijn/Utilitarian-distribution-of-OR-capacity-during-COVID-19.

## Results

### Data collection

In total, 12 cardiothoracic surgeries were evaluated, along with 23 oncological surgeries, 2 transplantations (liver and living donor kidney), 5 vascular surgeries, and 1 other type of surgery (creation of a shunt to facilitate haemodialysis). These 43 evaluated surgeries comprised of 69% of the total semi-elective program in our hospital.

Survival with treatment was mostly based on national registries (31/43). Survival without treatment was mostly based on data from (inter)national registries (12/43 surgeries, 6 indirectly calculated through the treatment effect), but also frequently from RCT’s (10/43 surgeries, 7 indirectly calculated), and observational studies (9/43 surgeries, 3 indirectly calculated). For 14/43 surgeries, QoL was available through the WHO Global Burden of Disease study.^26^ For the remaining 29 surgeries, the QoL of the pre- and postoperative health state was estimated by the expert panel as described in Appendix C. For 6/43 surgeries, a “time-to-no-effect-on-QoL” within one year, our maximum period of delaying surgery, was applicable. For 23 surgeries, a “time-to-no-effect-of-treatment-on-survival” was assumed based on qualitative assessment of the literature. Most of these surgeries were oncological surgeries (20/23). The estimates for the time until surgery becomes ineffective was mostly based on class IIb evidence (retrospective and prospective observational studies, see table 1). Input parameters varied widely between surgeries (Figure 2). Appendix A presents all input parameters, their sources^26,27,36–45,28,46–55,29,56–65,30,66–75,31,76–82,32–35^, and the corresponding model output for each semi-elective surgery.

**Table 1.**
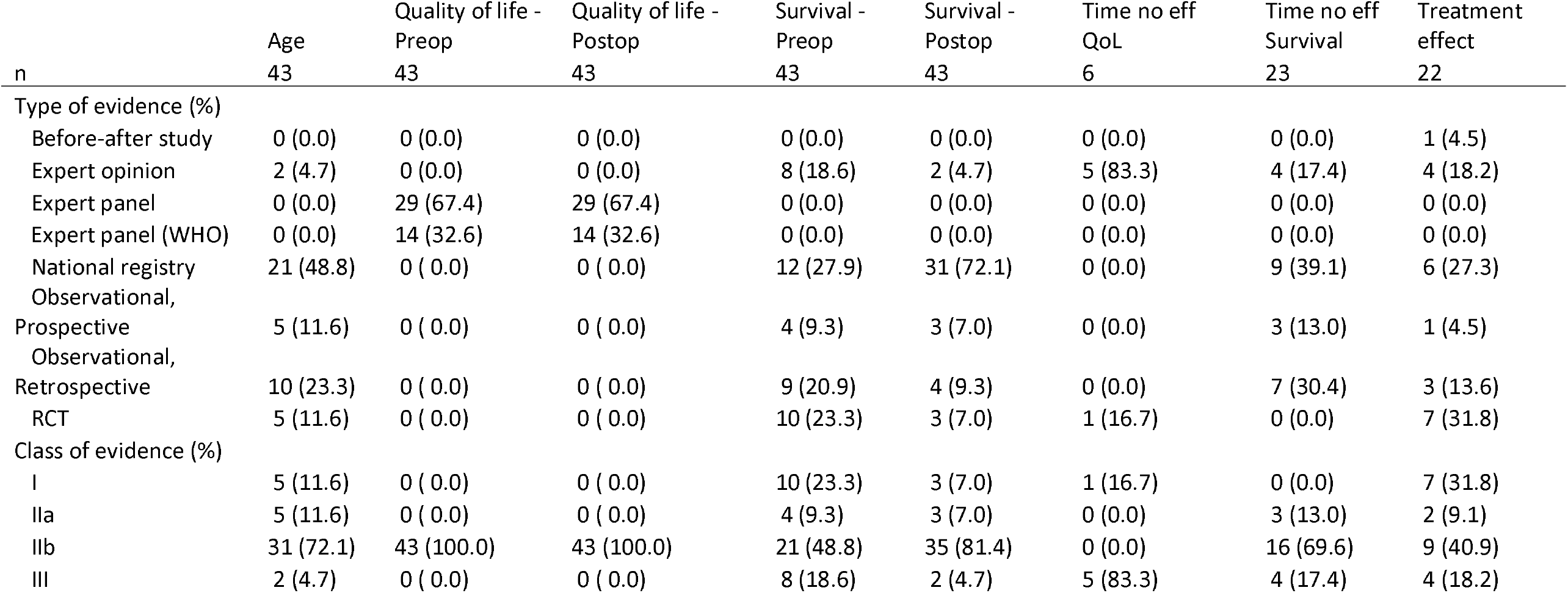
class and type of evidence underlying the model parameter inputs. Class definitions: I = Randomized Controlled Trials (RCT) or systematic reviews of RCTs, IIa = Prospective observational studies, before-after studies, IIb = Retrospective observational studies, expert panels for the utilities, national registries, class III = expert opinion. QoL = Quality of Life, Preop = preoperative, Postop = Postoperative.

| n | Age<br>43 | Quality of life -<br>Preop<br>43 | Quality of life -<br>Postop<br>43 | Survival -<br>Preop<br>43 | Survival -<br>Postop<br>43 | Time no eff<br>QoL<br>6 | Time no eff<br>Survival<br>23 | Treatment<br>effect<br>22 |
| --- | --- | --- | --- | --- | --- | --- | --- | --- |
| Type of evidence (%) |  |  |  |  |  |  |  |  |
| Before-after study | 0 (0.0) | 0 (0.0) | 0 (0.0) | 0 (0.0) | 0 (0.0) | 0 (0.0) | 0 (0.0) | 1 (4.5) |
| Expert opinion | 2 (4.7) | 0 (0.0) | 0 (0.0) | 8 (18.6) | 2 (4.7) | 5 (83.3) | 4 (17.4) | 4 (18.2) |
| Expert panel | 0 (0.0) | 29 (67.4) | 29 (67.4) | 0 (0.0) | 0 (0.0) | 0 (0.0) | 0 (0.0) | 0 (0.0) |
| Expert panel (WHO) | 0 (0.0) | 14 (32.6) | 14 (32.6) | 0 (0.0) | 0 (0.0) | 0 (0.0) | 0 (0.0) | 0 (0.0) |
| National registry | 21 (48.8) | 0 (0.0) | 0 (0.0) | 12 (27.9) | 31 (72.1) | 0 (0.0) | 9 (39.1) | 6 (27.3) |
| Observational, |  |  |  |  |  |  |  |  |
| Prospective | 5 (11.6) | 0 (0.0) | 0 (0.0) | 4 (9.3) | 3 (7.0) | 0 (0.0) | 3 (13.0) | 1 (4.5) |
| Observational, |  |  |  |  |  |  |  |  |
| Retrospective | 10 (23.3) | 0 (0.0) | 0 (0.0) | 9 (20.9) | 4 (9.3) | 0 (0.0) | 7 (30.4) | 3 (13.6) |
| RCT | 5 (11.6) | 0 (0.0) | 0 (0.0) | 10 (23.3) | 3 (7.0) | 1 (16.7) | 0 (0.0) | 7 (31.8) |
| Class of evidence (%) |  |  |  |  |  |  |  |  |
| I | 5 (11.6) | 0 (0.0) | 0 (0.0) | 10 (23.3) | 3 (7.0) | 1 (16.7) | 0 (0.0) | 7 (31.8) |
| IIa | 5 (11.6) | 0 (0.0) | 0 (0.0) | 4 (9.3) | 3 (7.0) | 0 (0.0) | 3 (13.0) | 2 (9.1) |
| IIb | 31 (72.1) | 43 (100.0) | 43 (100.0) | 21 (48.8) | 35 (81.4) | 0 (0.0) | 16 (69.6) | 9 (40.9) |
| III | 2 (4.7) | 0 (0.0) | 0 (0.0) | 8 (18.6) | 2 (4.7) | 5 (83.3) | 4 (17.4) | 4 (18.2) |

**Figure 2.**
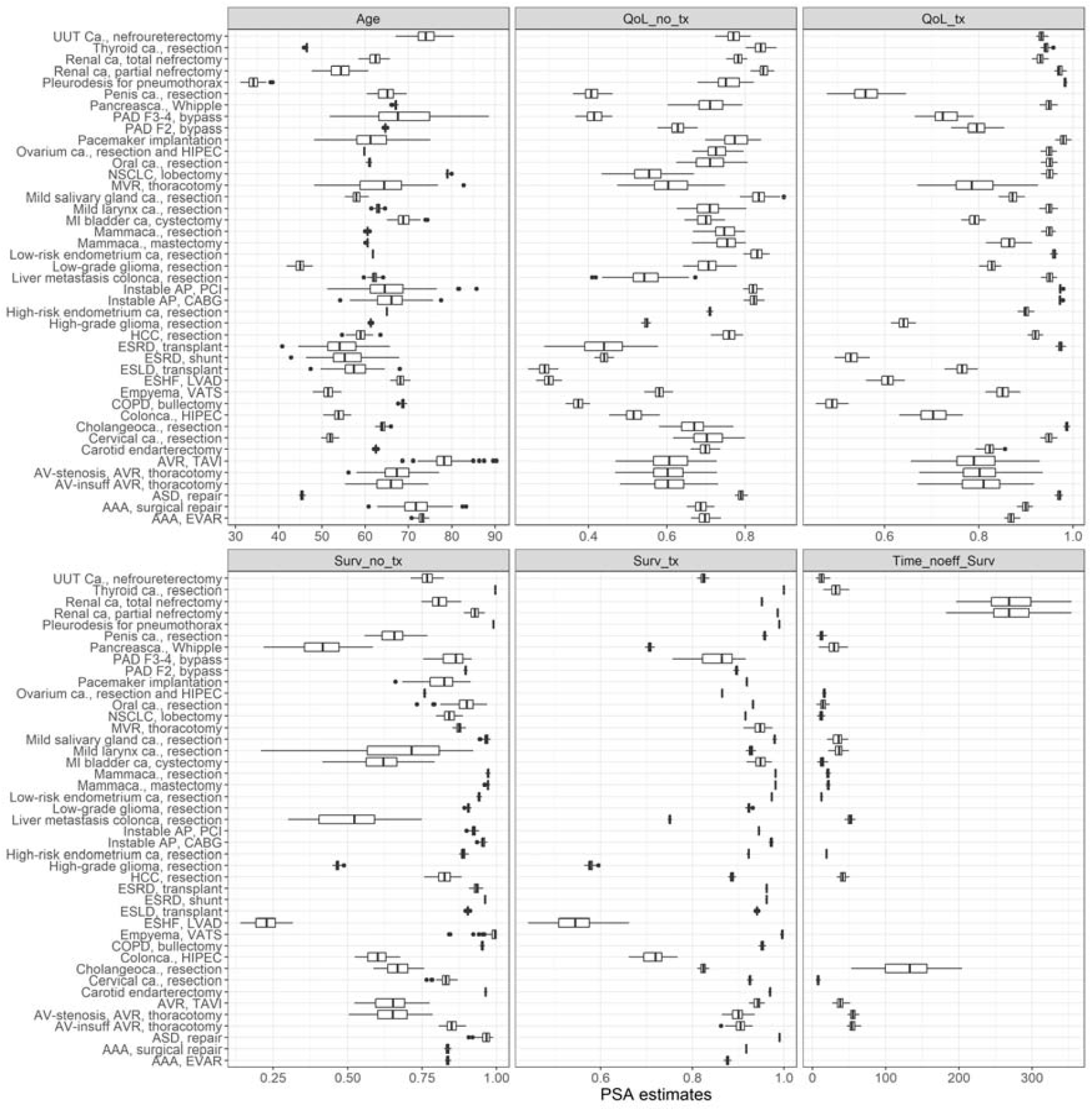
input parameters for the model. For a full list of input parameters per disease and source, see appendix A. **Abbreviations Figure titles:** Qol_no_tx: Quality of Life without treatment; QoL_tx: quality of life with treatment; Surv_no_tx: 1-year survival probability without treatment; Surv_tx: 1-year survival probability with treatment; Time_noeff_surv: days until no treatment is effective. **Abbreviations surgery/indications:** AAA: aneurysm of the abdominal aorta; AP: angina pectoris; AV: aortic valve; AVR: aortic valve replacement; ESRD: end-stage renal disease; ASD: atrial septum defect; -ca.: carcinoma; CABG: coronary artery bypass graft; COPD: chronic obstructive pulmonary disease; ESHF: end-stage heart failure; ESLD: end-stage liver disease; EVAR: endovascular aortic repair; HIPEC: hyperthermic intraperitoneal chemotherapy; HCC: hepatocellular carcinoma; NSCLC: non-small cell lung carcinoma; MVR: mitral valve replacement; PAD: peripheral arterial disease; PAD F2: peripheral arterial disease Fontaine classification 2, PCI: percutaneous coronary intervention; TAVI: transaortic valve implantation; UUT: upper urinary track; VATS: video assisted thoracoscopic surgery.

### Quality of Life

The preoperative and postoperative health state of 3 surgeries (one with a mild and severe subgroup) were estimated in both sessions, resulting in 8 double estimates of QoL. The gain in QoL due to surgery was consistent in the two sessions (the standardized mean difference was 0.025, 95% CI: −0.11 – 0.16, Appendix B: table 3 and figure 1).

The maximum expected benefit of surgery, i.e. in a scenario without delay, ranged from 0.48 QALYs (95% CI: 0.32 – 0.83) for resection of muscle invasive bladder cancer to 10.3 QALYs (95% CI: 8.7 – 11.9) for kidney transplantation (Figure 3). The ranking based on QALYs gained by surgery was correlated with the ranking based on LYs gained by surgery: The Spearman rank correlation coefficient between the ranking of surgeries based on LYs and QALYs was 0.45 (p=0.003).

**Figure 3.**
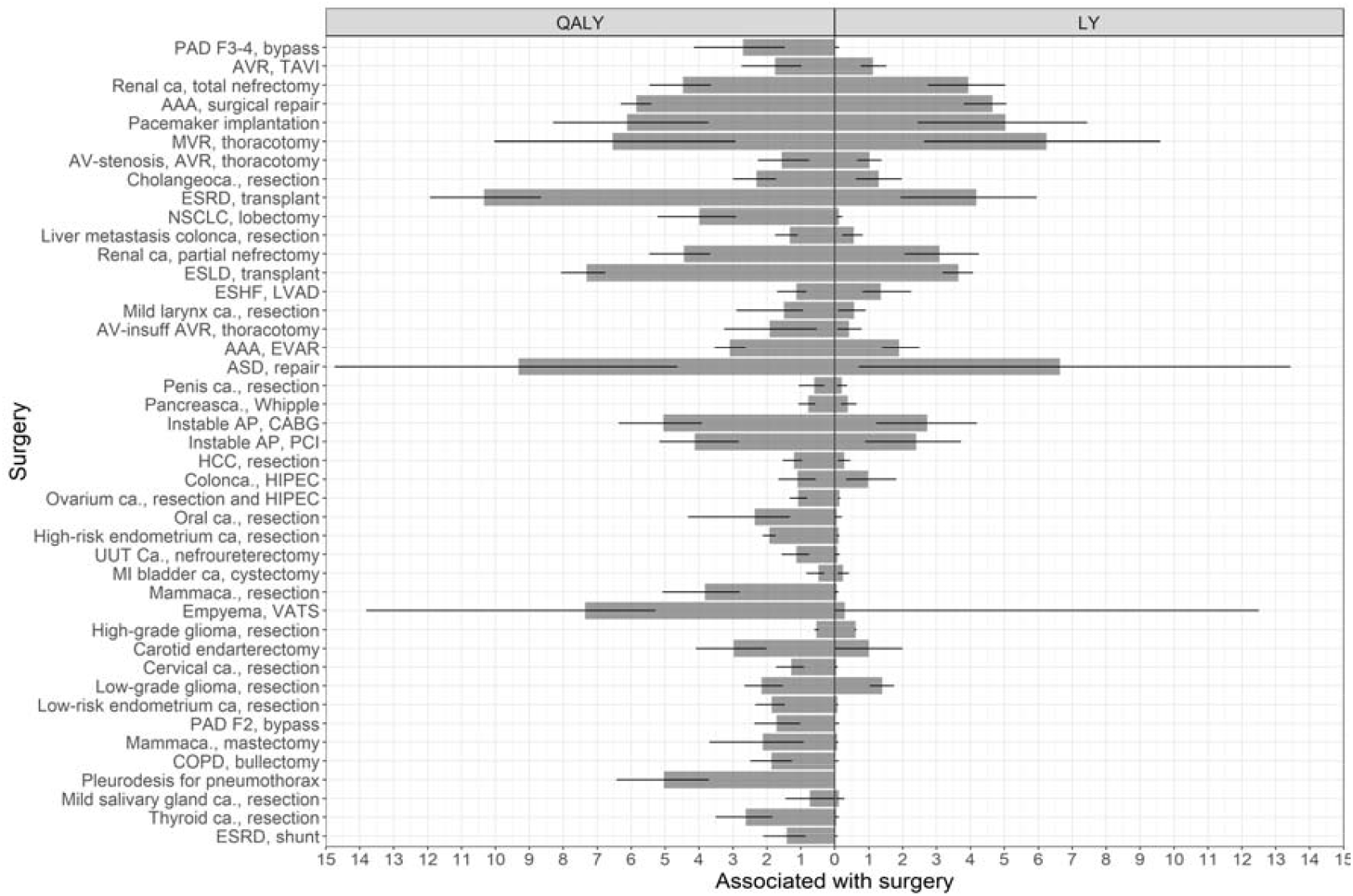
the maximum expected QALYs and LYs per surgery, in descending order of urgency (see figure 4). The estimates (gray bars) and 95% confidence intervals (black lines) are shown. The model output for no surgery was subtracted from the model output for a delay of 2 weeks. The actual data are presented in Appendix B. **Abbreviations Figure titles: QALY:** Quality of Life without treatment; LY: life years. **Abbreviations surgery/indication:** AAA: aneurysm of the abdominal aorta; AP: angina pectoris; AV: aortic valve; AVR: aortic valve replacement; ESRD: end-stage renal disease; ASD: atrial septum defect; - ca.: carcinoma; CABG: coronary artery bypass graft; COPD: chronic obstructive pulmonary disease; ESHF: end-stage heart failure; ESLD: end-stage liver disease; EVAR: endovascular aortic repair; HIPEC: hyperthermic intraperitoneal chemotherapy; HCC: hepatocellular carcinoma; NSCLC: non-small cell lung carcinoma; MVR: mitral valve replacement; PAD: peripheral arterial disease; PAD F2: peripheral arterial disease Fontaine classification 2, PCI: percutaneous coronary intervention; TAVI: transaortic valve implantation; UUT: upper urinary track; VATS: video assisted thoracoscopic surgery.

### Urgency

Most surgeries had a clear linear descend in terms of QALYs per delay, except surgeries where a time until no effect of treatment on survival was used (figure 1, appendix B).

The urgency of the surgeries ranged from 0.01 QALY loss/month (95% CI: 0.00-0.01) for placing a shunt for dialysis, to 0.23 QALY loss/month (0.09-0.24) for a bypass surgery for Fontaine III/IV peripheral arterial disease (Appendix B: figure 4 and table 1). If the latter would be postponed by a month, patients lose approximately 84 days (0.23*365) spent in perfect health of their remaining expected QALYs gained by surgery.

**Figure 4.**
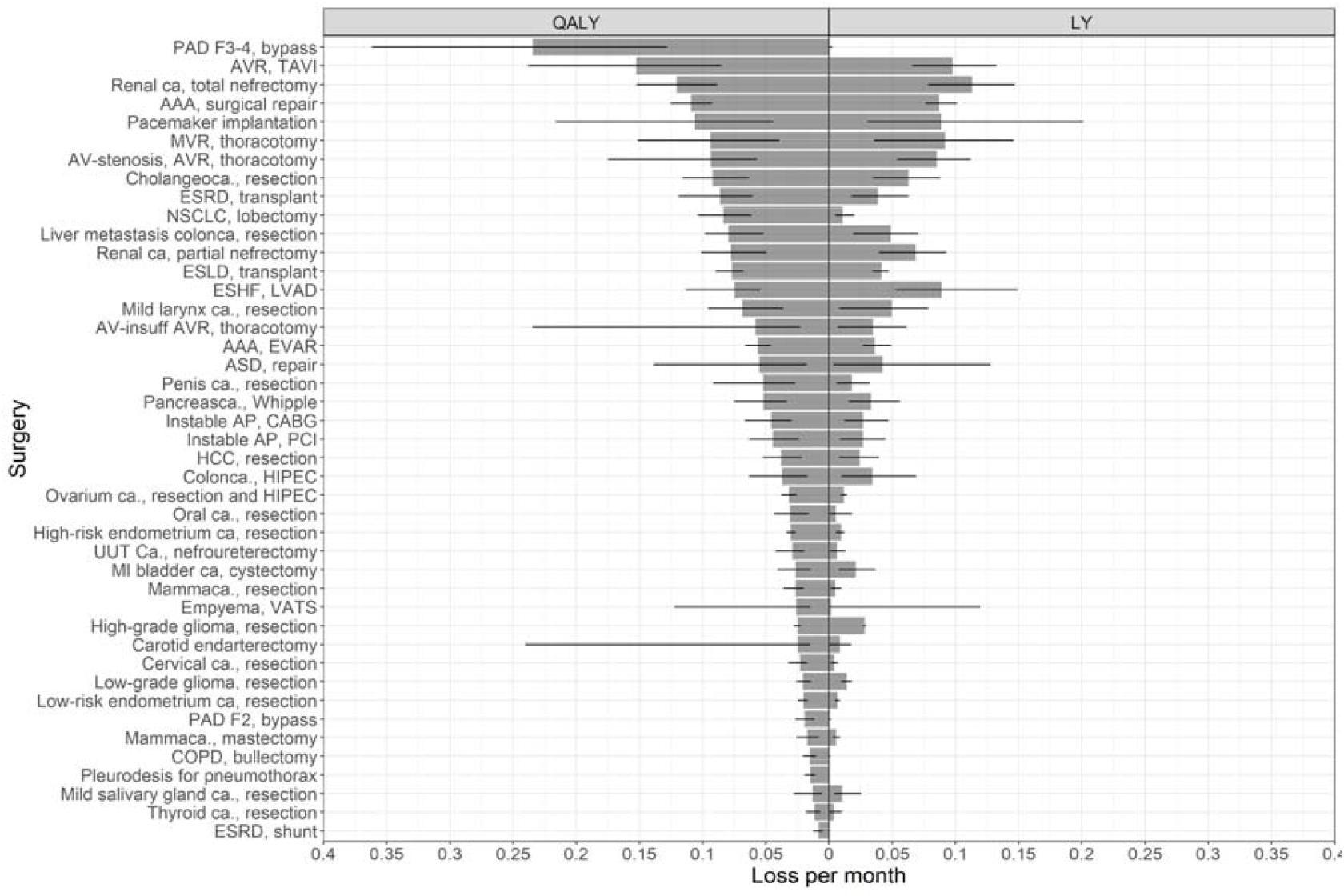
the average loss of QALYs and LYs per month of delay for the investigated surgeries based on the simulation of surgery delay of 52 weeks. The estimates (gray bars) and 95% confidence intervals (black lines) are shown. The actual data are presented in appendix B. **Abbreviations Figure titles:** QALY: Quality of Life without treatment; LY: life years. **Abbreviations diseases/indication:** AAA: aneurysm of the abdominal aorta; AP: angina pectoris; AV: aortic valve; AVR: aortic valve replacement; ESRD: end-stage renal disease; ASD: atrial septum defect; -ca.: carcinoma; CABG: coronary artery bypass graft; COPD: chronic obstructive pulmonary disease; ESHF: end-stage heart failure; ESLD: end-stage liver disease; EVAR: endovascular aortic repair; HIPEC: hyperthermic intraperitoneal chemotherapy; HCC: hepatocellular carcinoma; NSCLC: non-small cell lung carcinoma; MVR: mitral valve replacement; PAD: peripheral arterial disease; PAD F2: peripheral arterial disease Fontaine classification 2, PCI: percutaneous coronary intervention; TAVI: transaortic valve implantation; UUT: upper urinary track; VATS: video assisted thoracoscopic surgery.

The most urgent surgeries after bypass surgery for Fontaine III/IV peripheral arterial disease, was transaortic valve implantation (0.15 QALY loss/month, 95% CI: 0.09-0.24). After placing a shunt for patients with end-stage renal disease, the least urgent surgery was resection of thyroid cancer (0.01 QALY loss/month, 95% CI: 0.01-0.02) (Appendix B). Surgeries that were associated with a higher expected QALY benefit, often lost more QALYs per month delay: The Spearman correlation coefficient between the ranking of health benefit, in terms of QALYs, and urgency, in terms of QALY loss/month, was 0.32 (p=0.04).

Urgency was strongly correlated with the LYs gained per surgery: The Spearman rank correlation coefficient between the ranking of surgeries based on LYs and QALY loss/month was 0.79 (p< 0.001).

### Capacity

Surgeries that are ranked high in terms of urgency and had relative short surgery time compared to other surgeries include repair of atrial septum defects (surgery time: 74 min [IQR: 56-131], 0.06 QALY loss/month [95% CI: 0.02-0.14]), pacemaker implantations (115 min [82-154], 0.11 QALY loss/month [0.04-0.22]), resection of mild larynx carcinoma (70 min [38-109], 0.07 QALY loss/month[0.04-0.11]), and valve replacements (99 min [77-125]; mitral valve replacement: 0.09 QALY loss/month [0.04-0.15]; aortic valve replacement: 0.09 QALY loss/month [0.06-0.17]) (Figure 5). Liver transplant is relatively urgent but requires an exceptional amount of OR-time (875 min [797-957], 0.08 QALY loss/month [0.07-0.09]) (table 2 Appendix B).

**Figure 5.**
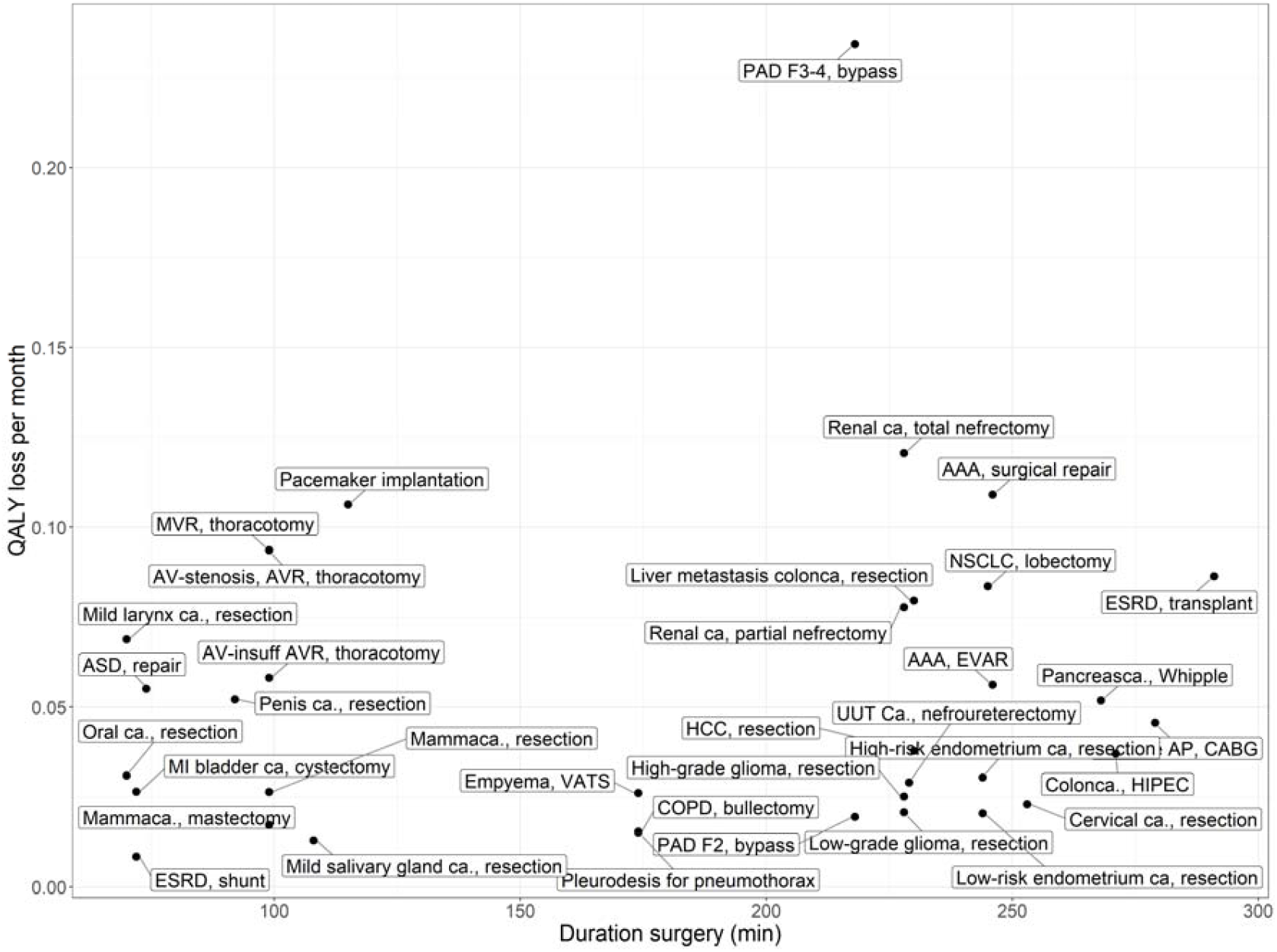
showing the mean duration of the surgeries and the urgency in terms of QALY loss per month. Liver transplant is excluded in this plot, because it was an outlier in terms of duration of surgeries (median: 875 minutes, IQR: 797-957 and - 0.08 QALY per month, 95% CI: −0.09--0.07). **Abbreviations Figure titles:** QALY: Quality of Life without treatment. **Abbreviations diseases/indication:** AAA: aneurysm of the abdominal aorta; AP: angina pectoris; AV: aortic valve; AVR: aortic valve replacement; ESRD: end-stage renal disease; ASD: atrial septum defect; -ca.: carcinoma; CABG: coronary artery bypass graft; COPD: chronic obstructive pulmonary disease; ESHF: end-stage heart failure; ESLD: end-stage liver disease; EVAR: endovascular aortic repair; HIPEC: hyperthermic intraperitoneal chemotherapy; HCC: hepatocellular carcinoma; NSCLC: non-small cell lung carcinoma; MVR: mitral valve replacement; PAD: peripheral arterial disease; PAD F2: peripheral arterial disease Fontaine classification 2, PCI: percutaneous coronary intervention; TAVI: transaortic valve implantation; UUT: upper urinary track; VATS: video assisted thoracoscopic surgery.

## Discussion

Our proposed decision model is an attempt to systematically guide prioritization of surgeries from a utilitarian perspective. Urgency was quantified based on the expected health loss due to surgery delay. Our approach operationalizes ethical values that are the most appropriate in times of scarcity.2 Available evidence suggests that semi-elective surgeries can be ranked based on their urgency using a simple decision model. For survival after surgery, most evidence was based on national registries, while treatment effects were mostly derived from RCTs. The time until no effect of treatment on survival or QoL, however, was most often class IIb/III evidence.

Among the 43 surgeries analysed, bypass surgery for Fontaine III/IV peripheral arterial disease and transaortic valve implantation were the most urgent surgeries. Less urgent surgeries were placement of a shunt for dialysis and resection of thyroid cancer. Liver transplantation shows to be a relatively urgent surgery but requires an exceptionally long surgery time. In times of scarce OR-capacity, this surgery is less efficient in the prevention of QALY loss.

Interestingly, the ranking of urgency is primarily driven by the gain in LY associated with surgery: Surgeries that are associated with substantial gain in LY (e.g. mitral valve replacement), also lose more QALYs per month delay than surgeries that are associated with no gain in LY (e.g. creation of a shunt for haemodialysis). The larger the total health benefit associated with surgery, the more health can potentially be lost by postponing surgery.

To optimize OR triage, our metric for urgency should be weighed against capacity indicators. This is effectively a cost-effectiveness analysis, where resource constraints represent costs. For the scenario where OR-capacity is the most scarce in terms of hospital capacity, urgency can be plotted against surgery time. This simple method revealed that repairing atrial septum defects, pacemaker implantations, resection of mild larynx carcinoma, and valve replacements are the most efficient surgeries in our hospital to perform in this context. However, there are contexts where other types of capacity (e.g. beds or staff) are scarcer, and therefore more relevant to be weighed against urgency.

Although our modelling approach rationalizes and objectively quantifies urgency from a utilitarian perspective, it needs to be complemented by other perspectives to be used effectively in practice. First, an important consideration from the medical perspective may be the availability of alternative treatment strategies. In cancer treatment, (chemo-)radiation or systematic therapy alone may be considered instead of surgery. Second, a financial perspective might also be explored. This perspective might be less relevant in a crisis such as the COVID-19 pandemic, where the bottleneck mainly seems hospital capacity instead of costs. If this approach would be applied to the context of regular care, this perspective might be of increasing importance. Finally, other ethical perspectives (e.g. rule of rescue^13^) might be explored to assess the viability of our approach, and it needs to be established whether our approach is applicable to all surgical procedures.

There are practical advantages of comparing “average patients” on urgency, despite the fact that there is no such thing as an “average patient”: It prevents our approach from systematically discriminating against a specific group of patients. Our approach would only discriminate if specific socioeconomic groups would suffer more frequently from diseases that are less urgent. It is known that lower socioeconomic groups are more prone to develop diseases that have clear association with unhealthy behaviour, such as lung cancer.^83^ However, these diseases do not systematically rank low in our approach. Comparing the average patients across specialties on urgency may not be a personalized approach, but it can be tailored to an individual’s context by providing input for shared decision making. We feel that next to a quantitative estimation of urgency from a utilitarian perspective, individual patient’s preferences, social contexts, and operability should also be included in the decision making process.

Since all models are a simplification of reality, our model has several limitations. First, the survival data used were not always derived from high-quality evidence. Although survival with treatment might be validly estimated from national registries, the survival without treatment is harder to be unbiasedly estimated. The surgeries that were evaluated are often part of standard clinical practice. Therefore, data might be biased (e.g. selection bias in the survival without treatment because patients opt for palliative care), or not available (it would be unethical now to perform RCTs evaluating surgery versus no surgery). Instead, best available evidence was used, which in part included evidence from more historical RCTs. As such, data might be biased, and as a result, so are the estimates from our model. Because of this limitation, our approach is simply to aggregate transparently and systematically the best currently available evidence using a model.

Second, it was assumed that all surgeries are successful. The current model does not simulate adverse events, like major bleedings or death due to surgery. Also, the potential reduction of QoL due to these adverse events were not incorporated, nor the QoL reduction of a temporary period of recovery after surgery. Because of these assumptions, the overall QALYs associated with the surgery should not be interpreted as an absolute estimate. They are the maximum possible QALYs that can be acquired by performing the surgery. However, these assumptions were considered reasonable to achieve the main goal of this study: when surgery without delay is compared to surgery with delay, the harm in both scenarios is similar and therefore cancel out.

Third, because the health loss in 50 weeks was simply converted to loss per month, a linear approximation was effectively used to quantify urgency by delaying surgery up to a year. Some surgeries did show a slightly steeper decrease in the period up to 32 weeks delay. The data needed to validly model this decay in QALYs per unit of time for all surgeries likely don’t exist: most of the estimates of time to no effect on survival were based on observational studies, which are likely biased. A more detailed approximation would be possible using a more individualized model which also models the natural grow of tumours, or aneurysms, and validly model the development of metastasis. It was not feasible to develop this for all evaluated surgeries. Instead, we opted for a more pragmatic approach.

Fourth, QoL weights were derived from expert-opinion. In this approach the patient is not involved. Experts interpret the health states and give weights, thereby our approach takes a societal perspective. There are also multiple methodological, ethical, and contextual disadvantages of using QALYs, but it should be noted that most of those discussion are more about utilitarian principles.^84^

Fifth, the potential impact on QoL of delaying a semi-elective surgery was not included. This impact might differ across surgeries. It might be hypothesized that surgeries performed after already a long disease history (e.g. kidney transplant) might have less “waiting time disutility” than recently diagnosed diseases (e.g. mammacarcinoma).

Part of the input parameters were based on national registry data, but a substantial amount of the input originated from various international sources. Therefore, with some modifications, the model can easily be adapted to different contexts. Therefore, this study can be considered the first step towards a triaging strategy which optimizes surgical benefit in times of scarcity in surgical capacity, such as during the COVID-19 pandemic. To improve validity, it is however essential to periodically review the literature and update the model with higher quality evidence, much like a living systematic review.^85^ If accepted, a wider range of surgeries should be considered, implementation strategies should be explored and evaluated, and the model should be applied to a variety of settings.

## Conclusion

By transparently aggregating best available evidence, our decision model support prioritization of surgical care in times of scarce surgical capacity (e.g. during pandemics) from a utilitarian perspective. Our approach quantifies the expected health loss due to delay for semi-elective surgeries in an academic hospital in the Netherlands. This approach can help to minimize health losses when trying to overcome delay in surgeries across disciplines. This approach is more transparent, more evidence-based, and more consistent than the alternative strategy of triaging based on expert opinion.

It should be noted that evidence from well-controlled comparison studies is often lacking. Instead, adjusted estimates from observational studies are often the best available evidence for benefit of surgery and the effects of delay on survival. Therefore, model inputs should be periodically updated with newer, higher quality evidence.

Finally, our approach should be placed in the context of other ethical perspectives and combined with capacity management tools. If accepted, we believe this tool should be implemented on a large scale, in order to minimize health loss of the accumulating group of patients awaiting surgery.

## Data Availability

All source data and code are made available in an online github repository.

https://github.com/bgravesteijn/Utilitarian-distribution-of-OR-capacity-during-COVID-19

## Authors contribution

**Benjamin Gravesteijn:** Conceptualization (Lead), Data curation (Lead), Formal analysis (Lead) Investigation (Lead), Methodology (Lead), Validation (Equal), Visualization (Lead), Writing-original draft (Lead)

**Eline Krijkamp**: Conceptualization (Lead), Data curation (Equal), Formal analysis (Lead), Investigation (Lead), Methodology (Lead), Validation (Supporting), Visualization (Lead), Writing-original draft (Lead)

**Jan Busschbach**: Conceptualization (Equal), Investigation (Equal), Methodology (Equal), Supervision (Supporting), Validation (Equal), Writing-review & editing (Equal)

**Geert Geleijnse**: Conceptualization (Equal), Data curation (Equal), Investigation (Supporting), Supervision (Supporting), Validation (Equal), Visualization (Supporting), Writing-review & editing (Equal)

**Isabel Retel Helmrich**: Data curation (Equal), Investigation (Equal), Methodology (Supporting), Writing-review & editing (Supporting)

**Sophie Bruinsma**: Data curation (Equal), Investigation (Equal), Writing-review & editing (Supporting)

**Céline van Lint**: Data curation (Equal), Investigation (Equal), Writing-review & editing (Supporting)

**Ernest van Veen**: Data curation (Supporting), Investigation (Supporting), Writing-review & editing (Supporting)

**Ewout Steyerberg**: Methodology (Supporting), Validation (Equal), Writing-review & editing (Supporting)

**Kees Verhoef**: Investigation (Supporting), Methodology (Supporting), Validation (Equal), Writing-review & editing (Supporting)

**Jan van Saase**: Conceptualization (Equal), Investigation (Equal), Supervision (Supporting), Validation (Equal), Writing-original draft (Equal), Writing-review & editing (Equal)

**Hester Lingsma**: Conceptualization (Lead), Investigation (Lead), Supervision (Lead), Writing-original draft (Equal), Writing-review & editing (Equal)

**Rob Baatenburg de Jong**: Conceptualization (Lead), Investigation (Lead), Supervision (Lead), Validation (Supporting), Writing-original draft (Equal), Writing-review & editing (Equal)

## Patient and public involvement

Already in the early phases, in the process of the design of this study, patient and public involvement was achieved by communicating our ideas via press-releases, interviews and open discussions. The aim for having this discussion was to identify barriers and facilitators of our approach, also possibly informing the study design.

Of course, it was not appropriate to include patient and/or public in the analysis of the study and writing the manuscript. However, medical experts, representing a part of the public, were included in the process of data collection for the quality of life values and further development of the methods. Moreover, the department of surgery within our hospital, an important end-user of our approach, tested the approach on face-validity and helped writing the manuscript, thereby showing their support.

Currently, we are also presenting our approach to patient federations. They have already expressed widespread interest and support to our approach. Finally, we are discussing with these stakeholders possible other barriers and facilitators for implementation which were not already identified.

## Acknowledgement

We are grateful for Lisa Caulley for her suggestions to revise the final manuscript. We are grateful for H. Karreman and C. Van der Velden-van der Graaf for the work they have done for the quality of life data collection. Moreover, we want to thank Ruben Goedhart, Esther van Spronsen and Linda van der Sluijs – van der Beek for extracting the data from the electronic patient registry.

## Appendix A

An overview per disease of the distribution and source of the input parameters and a graphical representation of the output of the model.

## Appendix B

A summary of the estimates of the decision model and an overview of the counts, duration, and length of stay of the included surgeries in our hospital.

## Appendix C

In-dept methods description.

## Footnotes

i A semi-elective surgery is defined as a surgery that should ideally be performed within three days up to three weeks.

## References

1 Office of the Assistant Secretary for Preparedness H. Pandemic Influenza Plan - Update IV (December 2017). 2017.

2 Emanuel EJ, Persad G, Upshur R, et al. Fair Allocation of Scarce Medical Resources in the Time of Covid-19. N Engl J Med 2020; : 1–7.

3 D’Agostino A, Demartini B, Cavallotti S, Gambini O. Mental health services in Italy during the COVID-19 outbreak. The Lancet Psychiatry. 2020; 7: 385–7.

4 Lazzerini M, Barbi E, Apicella A, Marchetti F, Cardinale F, Trobia G. Delayed access or provision of care in Italy resulting from fear of COVID-19. Lancet Child Adolesc. Heal. 2020; 4: e10–1.

5 Harahsheh AS, Dahdah N, Newburger JW, et al. Missed or Delayed Diagnosis of Kawasaki Disease During the 2019 Novel Coronavirus Disease (COVID-19) Pandemic. J Pediatr Pandemic J Pediatr 2020. DOI:10.1016/j.jpeds.2020.04.052.

6 Chang H-J, Huang N, Lee C-H, Hsu Y-J, Hsieh C-J, Chou Y-J. The Impact of the SARS Epidemic on the Utilization of Medical Services: SARS and the Fear of SARS. Am J Public Health 2004; 94: 562–4.

7 NZA. Analyse van de gevolgen van de coronacrisis voor de reguliere zorg. 2020 https://zorgdomein.com/media/documents/NZa-analyse_van_de_gevolgen_van_de_coronacrisis_voor_de_reguliere_zorg_pdf (accessed May 17, 2020).

8 Sud A, Jones M, Broggio J, et al. Collateral damage: the impact on outcomes from cancer surgery of the COVID-19 pandemic. Ann Oncol 2020; 13 19.

9 Powell SN, Mullen T, Young L, Heald D, Iv ETP. SARS-CoV-2 Impact on Elective Orthopaedic Surgery: Implications for Post-Pandemic Recovery. J Bone Jt Surg 2020. DOI:10.2106/JBJS.20.00690.

10 Salenger R, Etchill EW, Ad N, et al. The Surge after the Surge: Cardiac Surgery post-COVID-19.Ann Thorac Surg 2020; published online May 3. DOI:10.1016/j.athoracsur.2020.04.018.

11 Qadan M, Hong TS, Tanabe KK, Ryan DP, Lillemoe KD. A Multidisciplinary Team Approach for Triage of Elective Cancer Surgery at the Massachusetts General Hospital During the Novel Coronavirus COVID-19 Outbreak. Ann Surg 2020; 1.

12 MacCormick AD, Parry BR. Judgment analysis of surgeons’ prioritization of patients for elective general surgery. Med Decis Making 2006; 26: 255–64.

13 Garner RT, Rosen B. Moral Philosophy: A Systematic Introduction to Normative Ethics and Meta-Ethics. New York: Macmillan, 1967.

14 Vergano M, Bertolini G, Giannini A, et al. Clinical Ethics Recommendations for the Allocation of Intensive Care Treatments in exceptional, resource-limited circumstances. 2020 http://www.siaarti.it/SiteAssets/News/COVID19-documentiSIAARTI/SIAARTI-Covid-19ClinicalEthicsReccomendations.pdf (accessed May 17, 2020).

15 Daugherty Biddison L, Berkowitz KA, Courtney B, et al. Ethical considerations: Care of the critically ill and injured during pandemics and disasters: CHEST consensus statement. Chest 2014; 146: e145S–e155S.

16 Bayer R. Ethical Considerations for Decision Making Regarding Allocation of Mechanical Ventilators during a Severe Influenza Pandemic or Other Public Health Emergency 2011.

17 York State Department of Health N. VENTILATOR ALLOCATION GUIDELINES New York State Task Force on Life and the Law New York State Department of Health. 2015.

18 Toner E, Waldhorn R. Responding to pandemic influenza The ethical framework for policy and planning | Information | Health Service Journal 2020. https://www.hsj.co.uk/swine-flu/responding-to-pandemic-influenza-the-ethical-framework-for-policy-and-planning/5005219.article (accessed May 17, 2020).

19 Siebert U, Alagoz O, Bayoumi AM, et al. State-Transition Modeling: A Report of the ISPOR-SMDM Modeling Good Research Practices Task Force-3. Value Heal 2012; 15: 812–20.

20 Hunink M, Mc E, Glasziou P, Elstein A. Decision Making in Health and Medicine: Integrating Evidence and Values, 2nd edn. Cambridge: Cambridge University Press, 2003 http://www.cambridge.org (accessed May 19, 2020).

21 Klarman H, Rosenthal GD. Cost Effectiveness Analysis Applied to the Treatment of Chronic Renal Disease. Med Care 1968; 6.1: 48–54.

22 Sonnenberg FA, Beck JR. Markov Models in Medical Decision Making. Med Decis Mak 1993; 13: 322–38.

23 R Core Team. R: A language and Environment for Statistical Computing. 2013.

24 Alarid-Escudero F, Krijkamp EM, Enns EA, Hunink MGM, Pechlivanoglou P, Jalal H. Cohort state-transition models in R: From conceptualization to implementation. 2020; published online Jan 21. http://arxiv.org/abs/2001.07824 (accessed May 19, 2020).

25 Alarid-Escudero F, Krijkamp EM, Pechlivanoglou P, et al. A Need for Change! A Coding Framework for Improving Transparency in Decision Modeling. Pharmacoeconomics 2019; 37: 1329–39.

26 Salomon JA, Haagsma JA, Davis A, et al. Disability weights for the Global Burden of Disease 2013 study. 2015 www.thelancet.com/lancetgh (accessed May 14, 2020).

27 Wang J, Yan C, Fu A. A randomized clinical trial of comprehensive education and care program compared to basic care for reducing anxiety and depression and improving quality of life and survival in patients with hepatocellular carcinoma who underwent surgery. Medicine (Baltimore) 2019; 98: e17552.

28 Verwaal VJ, Bruin S, Boot H, Van Slooten G, Van Tinteren H. 8-Year follow-up of randomized trial: Cytoreduction and hyperthermic intraperitoneal chemotherapy versus systemic chemotherapy in patients with peritoneal carcinomatosis of colorectal cancer. Ann Surg Oncol 2008; 15: 2426–32.

29 Konstantinides S, Geibel A, Olschewski M, et al. A comparison of surgical and medical therapy for atrial septal defect in adults. N Engl J Med 1995; 333: 469–73.

30 Fein DA, Mendenhall WM, Parsons JT, et al. Carcinoma of the oral tongue: A comparison of results and complications of treatment with radiotherapy and/or surgery. Head Neck 1994; 16: 358–65.

31 Jakola AS, Myrmel KS, Kloster R, et al. Comparison of a strategy favoring early surgical resection vs a strategy favoring watchful waiting in low-grade gliomas. JAMA - J Am Med Assoc 2012; 308: 1881–8.

32 Haruna A, Muro S, Nakano Y, et al. CT scan findings of emphysema predict mortality in COPD. Chest 2010; 138: 635–40.

33 Ruys AT, Heuts SG, Rauws EA, Busch ORC, Gouma DJ, Van Gulik TM. Delay in surgical treatment of patients with hilar cholangiocarcinoma: Does time impact outcomes? HPB 2014; 16: 469–74.

34 Shin DW, Cho J, Kim SY, et al. Delay to curative surgery greater than 12 weeks is associated with increased mortality in patients with colorectal and breast cancer but not lung or thyroid cancer. Ann Surg Oncol 2013; 20: 2468–76.

35 Chen EY, Mayo SC, Sutton T, et al. Effect of Time to Surgery of Colorectal Liver Metastases on Survival. J Gastrointest Cancer 2020. DOI:10.1007/s12029-020-00372-5.

36 Yusuf S, Zucker D, Passamani E, et al. Effect of coronary artery bypass graft surgery on survival: overview of 10-year results from randomised trials by the Coronary Artery Bypass Graft Surgery Trialists Collaboration. Lancet 1994; 344: 563–70.

37 Noorbakhsh A, Tang JA, Marcus LP, et al. Gross-total resection outcomes in an elderly population with glioblastoma: A SEER-based analysis. Clinical article. J Neurosurg 2014; 120: 31–9.

38 Nakano R, Ohira M, Kobayashi T, et al. Hepatectomy versus stereotactic body radiotherapy for primary early hepatocellular carcinoma: A propensity-matched analysis in a single institution. Surg (United States) 2018; 164: 219–26.

39 Lee JN, Kwon SY, Choi GS, et al. Impact of surgical wait time on oncologic outcomes in upper urinary tract urothelial carcinoma. J Surg Oncol 2014; 110: 468–75.

40 Lim C, Bhangui P, Salloum C, et al. Impact of time to surgery in the outcome of patients with liver resection for BCLC 0-A stage hepatocellular carcinoma. J Hepatol 2018; 68: 100–8.

41 Moss AJ, Jackson Hall W, Cannom DS, et al. Improved survival with an implanted defibrillator in patients with coronary disease at high risk for ventricular arrhythmia. N Engl J Med 1996; 335: 1933–40.

42 Kankersoorten - IKNL. https://iknl.nl/kankersoorten (accessed May 19, 2020).

43 Scott SWM, Batchelder AJ, Kirkbride D, Naylor AR, Thompson JP. Late Survival in Nonoperated Patients with Infrarenal Abdominal Aortic Aneurysm. Eur J Vasc Endovasc Surg 2016; 52: 444–9.

44 Nyboe C, Karunanithi Z, Nielsen-Kudsk JE, Hjortdal VE. Long-term mortality in patients with atrial septal defect: a nationwide cohort-study. DOI:10.1093/eurheartj/ehx633.

45 Brewster DC, Jones JE, Chung TK, et al. Long-term outcomes after endovascular abdominal aortic aneurysm repair: The First Decade. Ann. Surg. 2006; 244: 426–36.

46 Brunner M, Olschewski M, Geibeli A, Bode C, Zehender M. Long-term survival after pacemaker implantation: Prognostic importance of gender and baseline patient characteristics. Eur Heart J 2004; 25: 88–95.

47 Rose EA, Gelijns AC, Moskowitz AJ, et al. Long-term use of a left ventricular assist device for end-stage heart failure. N Engl J Med 2001; 345: 1435–43.

48 Mazzone E, Preisser F, Nazzani S, et al. More Extensive Lymph Node Dissection Improves Survival Benefit of Radical Cystectomy in Metastatic Urothelial Carcinoma of the Bladder. Clin Genitourin Cancer 2019; 17: 105-113.e2.

49 Kann BH, Verma V, Stahl JM, et al. Multi-institutional analysis of stereotactic body radiation therapy for operable early-stage non-small cell lung carcinoma. Radiother Oncol 2019; 134: 44–9.

50 Huang CE, Yang YH, Chen WC, et al. Nephroureterectomy increase 5 year survival in patients on dialysis with upper urinary tract urothelial carcinoma. Oncotarget 2017; 8: 79876–83.

51 NHR. https://nederlandsehartregistratie.nl/ (accessed May 19, 2020).

52 Shalowitz DI, Epstein AJ, Ko EM, Giuntoli RL. Non-surgical management of ovarian cancer: Prevalence and implications. Gynecol Oncol 2016; 142: 30–7.

53 Pedregal-Mallo D, Sánchez Canteli M López F, Álvarez-Marcos C, Llorente JL, Rodrigo JP. Oncological and functional outcomes of transoral laser surgery for laryngeal carcinoma. Eur Arch Oto-Rhino-Laryngology 2018; 275: 2071–7.

54 Kim WR, Lake JR, Smith JM, et al. OPTN/SRTR 2016 Annual Data Report: Liver. Am J Transplant 2018; 18: 172–253.

55 Muluk SC, Muluk VS, Kelley ME, et al. Outcome events in patients with claudication: A 15-year study in 2777 patients. J Vasc Surg 2001; 33: 251–8.

56 Holtzman A, Morris CG, Amdur RJ, Dziegielewski PT, Boyce B, Mendenhall WM. Outcomes after primary or adjuvant radiotherapy for salivary gland carcinoma. Acta Oncol (Madr) 2017; 56: 484–9.

57 Murphy MM, Simons JP, Hill JS, et al. Pancreatic resection: A key component to reducing racial disparities in pancreatic adenocarcinoma. Cancer 2009; 115: 3979–90.

58 Mikkola R, Kelahaara J, Heikkinen J, Lahtinen J, Biancari F. Poor late survival after surgical treatment of pleural empyema. World J Surg 2010; 34: 266–71.

59 Keeley EC, Boura JA, Grines CL. Primary angioplasty versus intravenous thrombolytic therapy for acute myocardial infarction: a quantitative review of 23 randomised trials. Lancet 2003; 361: 13–20.

60 Piehler JM, Crichlow RW. Primary Carcinoma of the Gallbladder. Arch Surg 1977; 112: 26–30.

61 Warner L, Chudasama J, Kelly CG, et al. Radiotherapy versus open surgery versus endolaryngeal surgery (with or without laser) for early laryngeal squamous cell cancer.Cochrane Database Syst. Rev. 2014; 2014. DOI:10.1002/14651858.CD002027.pub2.

62 Warlow C, Farrell B, Fraser A, Sandercock P, Slattery J. Randomised trial of endarterectomy for recently symptomatic carotid stenosis: Final results of the MRC European Carotid Surgery Trial (ECST). Lancet 1998; 351: 1379–87.

63 Soran A, Ozmen V, Ozbas S, et al. Randomized Trial Comparing Resection of Primary Tumor with No Surgery in Stage IV Breast Cancer at Presentation: Protocol MF07-01. Ann Surg Oncol 2018; 25: 3141–9.

64 Ginsberg RJ, Rubinstein L V. Randomized trial of lobectomy versus limited resection for T1 N0 non-small cell lung cancer. Ann Thorac Surg 1995; 60: 615–23.

65 Redden MD, Chin TY, van Driel ML. Surgical versus non-surgical management for pleural empyema. Cochrane Database Syst. Rev. 2017; 2017. DOI:10.1002/14651858.CD010651.pub2.

66 Sørensen VR, Heaf J, Wehberg S, Sørensen SS. Survival Benefit in Renal Transplantation Despite High Comorbidity. Transplantation 2016; 100: 2160–7.

67 Shalowitz DI, Epstein AJ, Buckingham L, Ko EM, Giuntoli RL. Survival implications of time to surgical treatment of endometrial cancers. Am J Obstet Gynecol 2017; 216: 268.e1-268.e18.

68 van Harten M, de Ridder M, Hamming-Vrieze O, Smeele L, Balm A, van den Brekel M. The association of treatment delay and prognosis in head and neck squamous cell carcinoma (HNSCC) patients in a Dutch comprehensive cancer center. Oral Oncol 2014; 50: 282–90.

69 Stewart JM, Tone AA, Jiang H, et al. The optimal time for surgery in women with serous ovarian cancer. Can J Surg 2016; 59: 223–32.

70 Davies L, Welch G. Thyroid cancer survival in the United States: Observational data from 1973 to 2005. Arch Otolaryngol - Head Neck Surg 2010; 136: 440–4.

71 Bleicher RJ, Ruth K, Sigurdson ER, et al. Time to surgery and breast cancer survival in the United States. JAMA Oncol 2016; 2: 330–9.

72 Morse E, Fujiwara RJT, Judson B, Mehra S. Treatment Times in Salivary Gland Cancer: National Patterns and Association with Survival. Otolaryngol - Head Neck Surg (United States) 2018; 159: 283–92.

73 USRDS. https://www.usrds.org/2015/view/ (accessed May 19, 2020).

74 Kirkegård J, Mortensen FV, Hansen CP, Mortensen MB, Sall M, Fristrup C. Waiting time to surgery and pancreatic cancer survival: A nationwide population-based cohort study. Eur J Surg Oncol 2019; 45: 1901–5.

75 Chung JH, Lee SH, Kim KT, Jung JS, Son HS, Sun K. Optimal Timing of Thoracoscopic Drainage and Decortication for Empyema. Ann Thorac Surg 2014; 97: 224–9.

76 Organ Procurement and Transplantation Network. https://optn.transplant.hrsa.gov/data/view-data-reports/national-data/ (accessed May 19, 2020).

77 Howard DPJ, Banerjee A, Fairhead JF, Hands L, Silver LE, Rothwell PM. Population-Based Study of Incidence, Risk Factors, Outcome, and Prognosis of Ischemic Peripheral Arterial Events: Implications for Prevention. Circulation 2015; 132: 1805–15.

78 Bonow RO, Greenland P. Population-wide trends in aortic stenosis incidence and outcomes. Circulation. 2015; 131: 969–71.

79 Badalato GM, Gaya JM, Hruby G, et al. Immediate radical cystectomy vs conservative management for high grade cT1 bladder cancer: Is there a survival difference? BJU Int 2012; 110: 1471–7.

80 Lee CT, Madii R, Daignault S, et al. Cystectomy delay more than 3 months from initial bladder cancer diagnosis results in decreased disease specific and overall survival. J Urol 2006; 175: 1262–7.

81 Tan WS, Trinh QD, Hayn MH, et al. Delayed nephrectomy has comparable long-term overall survival to immediate nephrectomy for cT1a renal cell carcinoma: A population-based analysis. Urol Oncol Semin Orig Investig 2020; 38: 74.e13-74.e20.

82 Janssen MWW, Linxweiler J, Terwey S, et al. Survival outcomes in patients with large (7cm) clear cell renal cell carcinomas treated with nephron-sparing surgery versus radical nephrectomy: Results of a multicenter cohort with long-term follow-up. PLoS One 2018; 13. DOI:10.1371/journal.pone.0196427.

83 Ae LXC, Reichman ME, Ae BAM, et al. Impact of socioeconomic status on cancer incidence and stage at diagnosis: selected findings from the surveillance, epidemiology, and end results: National Longitudinal Mortality Study. Cancer Causes Control 2009; 20: 417–35.

84 Pettitt D, Raza S, Naughton B, et al. The Limitations of QALY: A Literature Review. J Stem Cell Res Ther 2016; 6. DOI:10.4172/2157-7633.1000334.

85 Elliott JH, Synnot A, Turner T, et al. Living systematic review: 1. Introductiondthe why, what, when, and how on behalf of the Living Systematic Review Network. DOI:10.1016/j.jclinepi.2017.08.010.

